# Epidemiology of Long-Term Catheter Implantation for Chemotherapy in the Public Health System in Brazil: A 16-Year Analysis of 168,869 Cases

**DOI:** 10.1101/2025.11.05.25339622

**Authors:** Arthur Soares Freitas e Silva, Nahmam Jarouche, Carolina Carvalho Jansen Sorbello, Bruno Jeronimo Ponte, Marcelo Fiorelli Alexandrino da Silva, Marcelo Passos Teivelis, Nelson Wolosker

**Affiliations:** Hospital Israelita Albert Einstein, São Paulo, SP, Brasil; Universidade de São Paulo - USP, Faculdade de Medicina, Departamento de Cirurgia Vascular e Endovascular, São Paulo, SP, Brasil; Faculdade Israelita de Ciências da Saúde Albert Einstein, São Paulo, SP, Brasil

**Keywords:** [Central Venous Cathethers], [Big Data], [Chemotherapy], [Unified Health System], [Implantable Catheters]

## Abstract

**Background:** The implantation of long term catheters is crucial in various clinical contexts, particularly for patients requiring prolonged intravenous systemic therapy for cancer treatment. Consequently, analyzing the epidemiology of long term catheter implantation is highly relevant to clinical practice and the management of public health resources, especially concerning the entire population of a country.

**Objectives:** To evaluate the placement of 168,869 long term chemotherapy catheters performed within Brazil public health system (SUS) from 2008 to 2023.

**Methods:** We used data from DATASUS (Department of Information and Informatics of the Brazilian Public Health System). Our analysis focused on patients treated in Brazil from 2008 to 2023. The study also analyzed patient demographic and clinical characteristics, number of procedures over the years and divided by macro-region, days of hospitalization and associated diagnoses, as well as mortality rates and financial transfers related to catheter placement.

**Results:** A total of 168,869 long term catheter implants were carried out, with over 70% of these procedures being performed in the South and Southeastern regions of the country. Most of the surgical interventions were performed on female patients (59.7%) and adult patients aged 30 to 64 years (57.4%). The predominant indications were neoplasms (78.9%), particularly colorectal (22.3%) and breast (19.4%). The average reimbursement to the SUS was US$115.33 per procedure, with a cumulative total of US$19.48 million in the period analyzed, and the mortality was 13 deaths per 1,000 catheter implants.

**Conclusions:** Based on a total of 168,869 chemotherapy catheters implanted from 2008 to 2023 in the Brazilian public health system, we observed that most of the patients were adult women with neoplasms. The associated mortality rate is low (1.3%) and the cost estimated per procedure is US$115.33.

## INTRODUCTION

The selection of a long-term central venous access catheter is crucial for the therapeutic planning of cancer patients undergoing chemotherapy, as it directly impacts the quality of medical care [1]. This choice depends on the clinical context, the preferences of patients, and the alignment of the team responsible for their care [2].

The most commonly used catheters for chemotherapy infusion include semi-implantable catheters, such as Hickman, Permcath and PICC (Peripherally Inserted Central Venous Catheter) [3], as well as fully implantable catheters (FIC), also known as portocaths [4], which are inserted through a peripheral or central vein. They are connected to a reservoir that is implanted over the muscle fascia at the chosen site. Unlike semi-implantable models, fully implantable catheters do not have an externalised segment [5], which reduces the risk of infection [6] and enhances durability compared to semi-implantable catheters [7].

Typically, the preferred sites for insertion are the internal jugular, external jugular and femoral veins, femoral vein [8] and the brachial vein [9,10], especially in cancer patients [11].

In the United States, at least 7 million central venous catheters are inserted annually, with the global figure exceeding 10 million [12]. In 2001, around 500,000 long-term catheters were inserted annually in the United States [13]. However, there is currently no updated data available in the literature regarding the insertion rate of long-term catheters.

In Brazil, the largest country in Latin America, the public health system (SUS) provides universal, free and comprehensive healthcare to everyone within its territory, regardless of nationality or socioeconomic status. It is one of the largest and most complex public health systems in the world, ensuring the right to complete, universal and free access to healthcare for over 200 million people [14]. Of this population, at least 160 million depend solely on the SUS, meaning they do not have private healthcare plans [15].

There are currently no articles that analyze the implantation of long-term catheters among all individuals in the population of a country, especially in developing countries like Brazil.

This study aimed to evaluate the implantation of 168,869 long-term catheters for chemotherapy carried out in the Brazilian public health system (SUS) between 2008 and 2023. We used publicly available data obtained from DATASUS (SUS Department of Information and Informatics), considering patient demographic and clinical data, number of procedures over the years and their distribution by macro-region, length of hospitalization and associated diagnoses, as well as lethality and financial transfers related to catheter implantation.

## METHODS

This was a cross-sectional, retrospective population-based study that was approved by the institution’s Research Ethics Committee (CAAAE 35826320.2.0000.0071). As DATASUS is a public repository of anonymized data, informed consent was not required [15].

The data was extracted from DATASUS, the online database of the Unified Health System (SUS), which compiles information on government-funded hospitalizations across the country. The focus was on implantation procedures for semi- or totally-implantable long-term catheters carried out between 2008 and 2023. Data extraction was automated using a Python-based protocol (v. 2.7.13; Beaverton, OR, USA) developed by our IT department, running on Windows 10. The Selenium WebDriver (v. 3.1.8; Selenium HQ) and Pandas (v. 2.7.13; Lambda Foundry, Inc. and PyData Development Team, NY, USA) tools were used for data segregation and refinement.

The average Brazilian population from 2008 to 2022 was 196,918,278 [12]. Considering that 71% of the population uses exclusively the Brazilian public health service, this study’s population sample was approximately 140,000,000 patients [14].

Data related to the implantation of long-term catheters for chemotherapy was identified using DATASUS codes 04.06.02.007-8 (implantation of semi- or totally-implantable long-term catheter) and 04.06.02.062-0 (removal of semi- or totally-implantable long-term catheter) filtered by ICD-10 codes C00-C97 and D00-D48 related to neoplasms diagnosis.

Data compilation and tabulation were performed in .csv format using Microsoft Office Excel 2019 (Redmond, WA, USA). Population data by age group were obtained from the Brazilian Institute of Geography and Statistics (IBGE) [16]. Statistical analyses were based on the population relying solely on the Brazilian public health system, which is approximately 140,000,000 patients [17].

The total number of procedures was analyzed sequentially, followed by the number of procedures per macro-region of the country, the demographic profile of the patients, the distribution by age group and biological sex, the related diagnoses, the length of stay in the ward and ICU, the associated lethality and, finally, the amounts transferred.

There were studied the annual procedure counts, regional distributions, patient demographics (age and biological sex), length of hospital stay, ICU duration, lethality and financial reimbursements, and financial data in Brazilian Reais (R$) that was converted to US dollars (U$) by the quotation at the time of analysis (U$1 = R$5.46).

Statistical analyses were conducted using SPSS version 20.0 for Windows (IBM Corp, Armonk, NY). Linear regression was employed to analyze trends in long-term catheters for chemotherapy implantation across various age groups. Chi-square and likelihood ratio tests were used to evaluate differences in rates and lethality across the country’s macro-regions. A significance level of P ≤ 0.05 was set for all statistical tests.

## RESULTS

Between 2008 and 2023, 168,869 long-term catheter implants were performed in Brazil’s public health system.

Table 1 presents the annual number of catheter implants from 2008 to 2023, along with each year’s percentage share of the total. An increase was observed over the years, with 2,764 in 2008 and 1,774 in 2023, an estimated increase of 10.1% over the years (95% CI 10.0% to 10.2%; p-value <0.001).

**Table 1.** Number of Catheter Implants per year.

| YEAR | NUMBER OF CATHETER IMPLANTS | PERCENTAGE |
| --- | --- | --- |
| 2008 | 2,764 | 1,6% |
| 2009 | 4,149 | 2,5% |
| 2010 | 4,912 | 2,9% |
| 2011 | 6,095 | 3,6% |
| 2012 | 7,264 | 4,3% |
| 2013 | 8,415 | 5,0% |
| 2014 | 9,363 | 5,5% |
| 2015 | 10,951 | 6,5% |
| 2016 | 11,392 | 6,7% |
| 2017 | 12,032 | 7,1% |
| 2018 | 13,024 | 7,7% |
| 2019 | 13,797 | 8,2% |
| 2020 | 14,196 | 8,4% |
| 2021 | 15,160 | 9,0% |
| 2022 | 16,661 | 9,8% |
| 2023 | 18,744 | 11,1% |
| TOTAL | 168,869 | 100% |

Table 2 presents the regional distribution of long-term catheter implant procedures in Brazil from 2008 to 2023. The Southeast region accounted for the largest share (44.1 %), followed by the South (29.5 %). The North contributed the fewest with 3,114 (1.8 %). When adjusting for population, the South leads with an implant rate of 1,163 per million inhabitants, followed by the Southeast at 877, and the North at only 180.

**Table 2.**
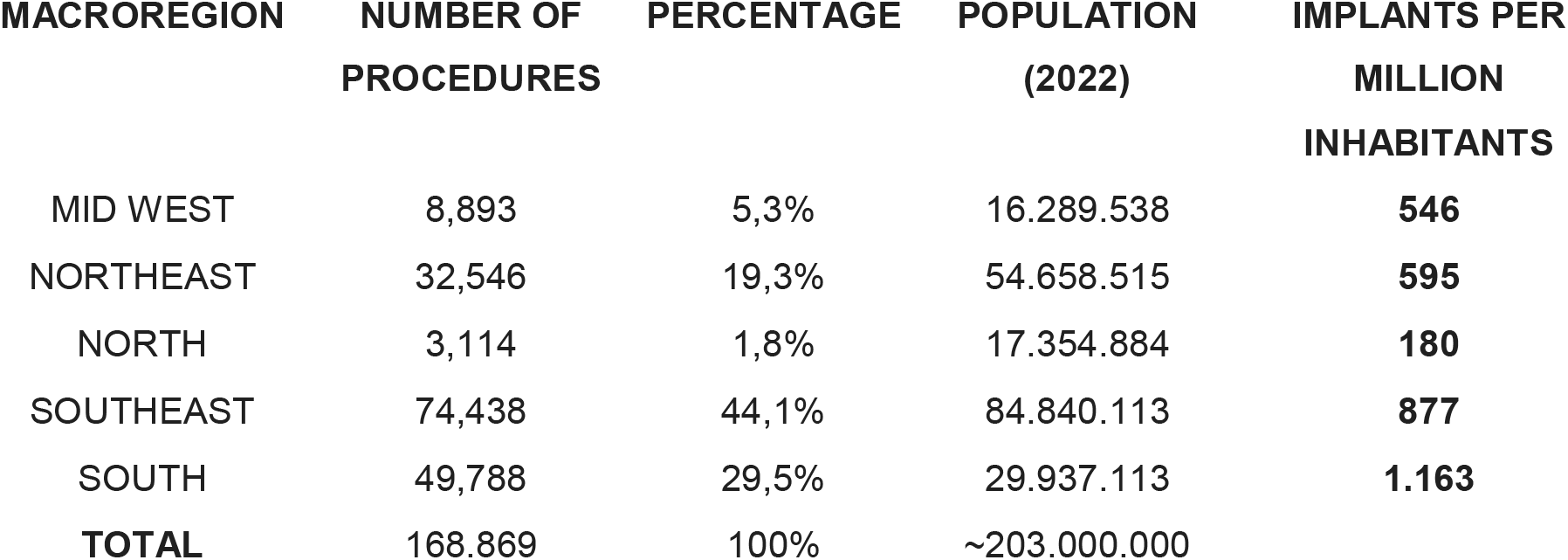
Distribution of Long-Term Catheter Implants by macroregion (2008-2023)

| MACROREGION | NUMBER OF<br>PROCEDURES | PERCENTAGE | POPULATION<br>(2022) | IMPLANTS PER<br>MILLION<br>INHABITANTS |
| --- | --- | --- | --- | --- |
| MID WEST | 8,893 | 5,3% | 16.289.538 | **546** |
| NORTHEAST | 32,546 | 19,3% | 54.658.515 | **595** |
| NORTH | 3,114 | 1,8% | 17.354.884 | **180** |
| SOUTHEAST | 74,438 | 44,1% | 84.840.113 | **877** |
| SOUTH | 49,788 | 29,5% | 29.937.113 | **1.163** |
| **TOTAL** | 168.869 | 100% | ~203.000.000 |  |

Table 3 presents the distribution of long-term catheter implant procedures in Brazil by age group, along with regional population and implant rate per million. Most procedures (57.5 %) were performed in adults aged 30–64 years. However, when adjusted for the implant rates per million inhabitants, seniors were the greatest users (1,618 per million inhabitants), followed by the pediatric group (574 per million).

**Table 3.** Distribution of Long-Term Catheter implants per age group (2008-2023)

| AGE GROUP | NUMBER OF<br>PROCEDURES | PERCENTAGE<br>OF TOTAL | POPULATION<br>(2022) | IMPLANTS<br>PER MILLION<br>INHABITANTS |
| --- | --- | --- | --- | --- |
| PEDIATRIC (0-14 YEARS) | 23,052 | 13,6% | 40.129.261 | **574** |
| ADOLESCENTS AND YOUNG<br>ADULTS (15-29 YEARS) | 12,913 | 7,6% | 45.312.128 | **285** |
| ADULTS (30-64 YEARS) | 97,031 | 57,5% | 95.470.266 | **1016** |
| SENIORS (65 YEARS AND<br>OLDER) | 35,873 | 21,2% | 22.169.101 | **1.618** |
| **TOTAL** | 168.869 | 100% | 203.080.756 |  |

Statistically significant differences were identified between age categories (p < 0.001): considering pediatrics as the reference, the 15–29 age group had a rate corresponding to 49.6% of pediatric values, adults (30–64 years) showed a 1.77-fold increase, and seniors a 2.82-fold higher rate.

Pediatric age group (0-14 years); Adolescents and young people (15-29 years); Adults (30-64 years); Seniors (65 years and older).

Figure 1 presents the analysis of the gender profile. Of 168,869 implants, 100,736 were in women (59.7%), with no change in sex distribution over the years (p-value >0.99). The number of procedures in women is estimated to be 1.48 times the number of procedures in men (95% CI 1.46 to 1.49; p-value <0.001).

**Figure 1.**
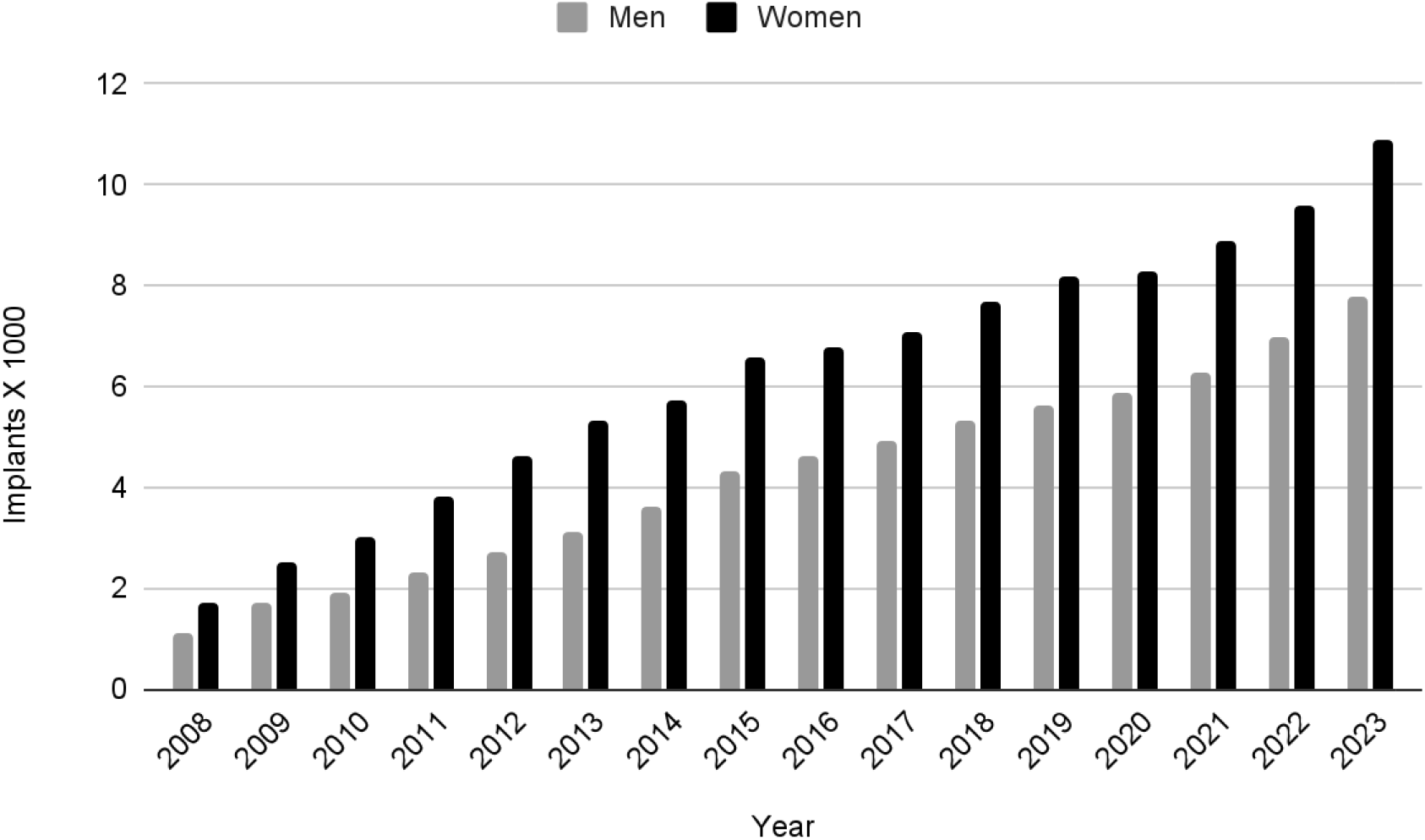
Analysis of the demographic profile.

Table 4 presents the primary diagnoses associated with long-term catheter implant procedures. The distribution underscores that a large proportion of catheter implants were linked to cancer-related vascular access needs (78.9%). Digestive tract neoplasms account for the largest share (22.31 %), followed by neoplasms of the breast (15.34 %).

**Table 4.**
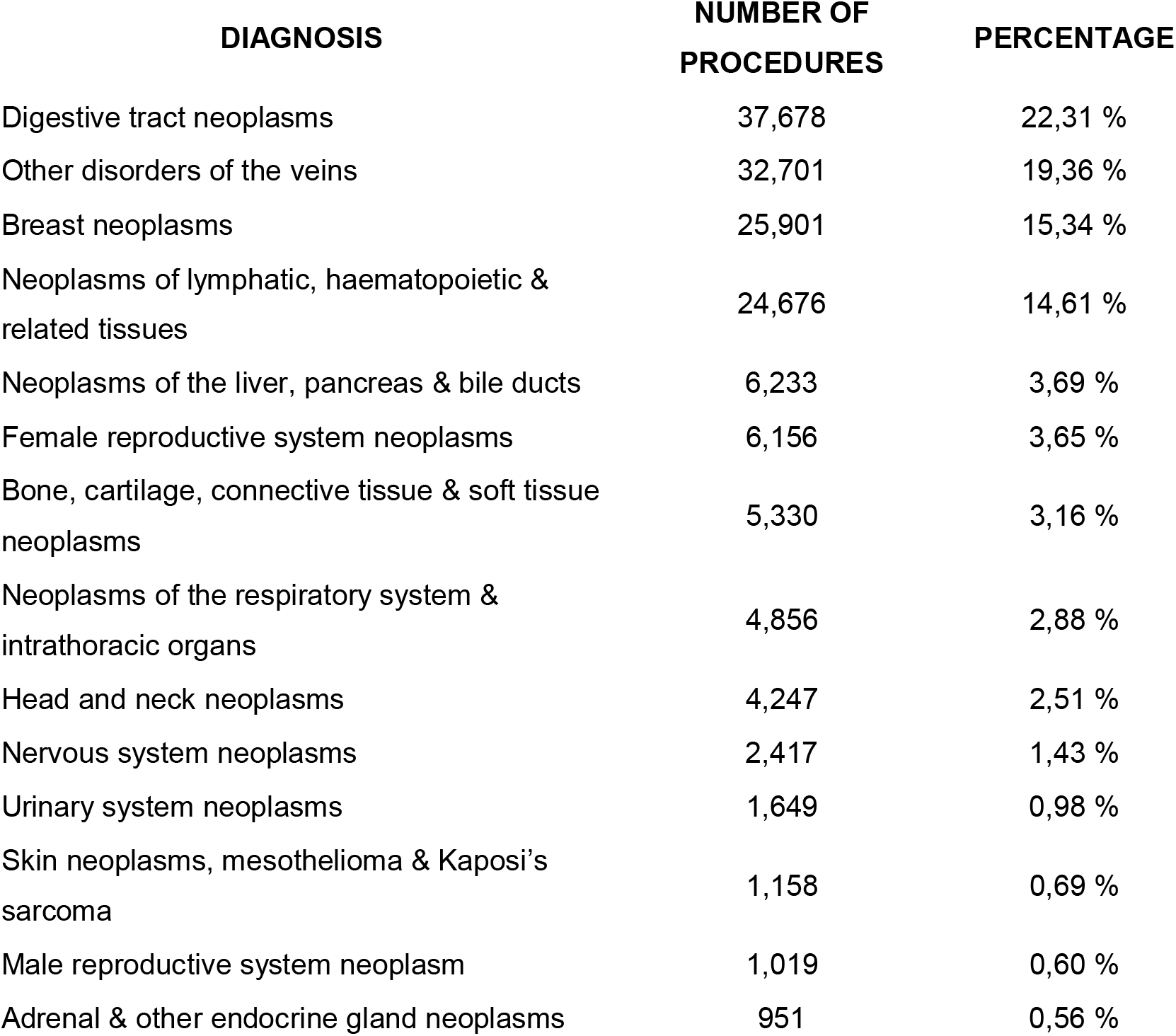
Diagnosis related to long-term catheter implantation (2008-2023).

| DIAGNOSIS | NUMBER OF<br>PROCEDURES | PERCENTAGE |
| --- | --- | --- |
| Digestive tract neoplasms | 37,678 | 22,31 % |
| Other disorders of the veins | 32,701 | 19,36 % |
| Breast neoplasms | 25,901 | 15,34 % |
| Neoplasms of lymphatic, haematopoietic &<br>related tissues | 24,676 | 14,61 % |
| Neoplasms of the liver, pancreas & bile ducts | 6,233 | 3,69 % |
| Female reproductive system neoplasms | 6,156 | 3,65 % |
| Bone, cartilage, connective tissue & soft tissue<br>neoplasms | 5,330 | 3,16 % |
| Neoplasms of the respiratory system &<br>intrathoracic organs | 4,856 | 2,88 % |
| Head and neck neoplasms | 4,247 | 2,51 % |
| Nervous system neoplasms | 2,417 | 1,43 % |
| Urinary system neoplasms | 1,649 | 0,98 % |
| Skin neoplasms, mesothelioma & Kaposi's<br>sarcoma | 1,158 | 0,69 % |
| Male reproductive system neoplasm | 1,019 | 0,60 % |
| Adrenal & other endocrine gland neoplasms | 951 | 0,56 % |

Most patients (74.3 %) remained hospitalized for no more than one day, and nearly half (46.3 %) were discharged on the same day.

Figure 2 illustrates the temporal trend of lethality over the years in Brazil. During the study period, 2,217 deaths were reported, corresponding to 13 deaths per 1,000 catheter implantations. Mortality rates ranged from 8 to 35 per 1,000 procedures, but showed a significant annual decline of 5.3% (95% CI: 4.4-6.2; p <0.001).

**Figure 2.**
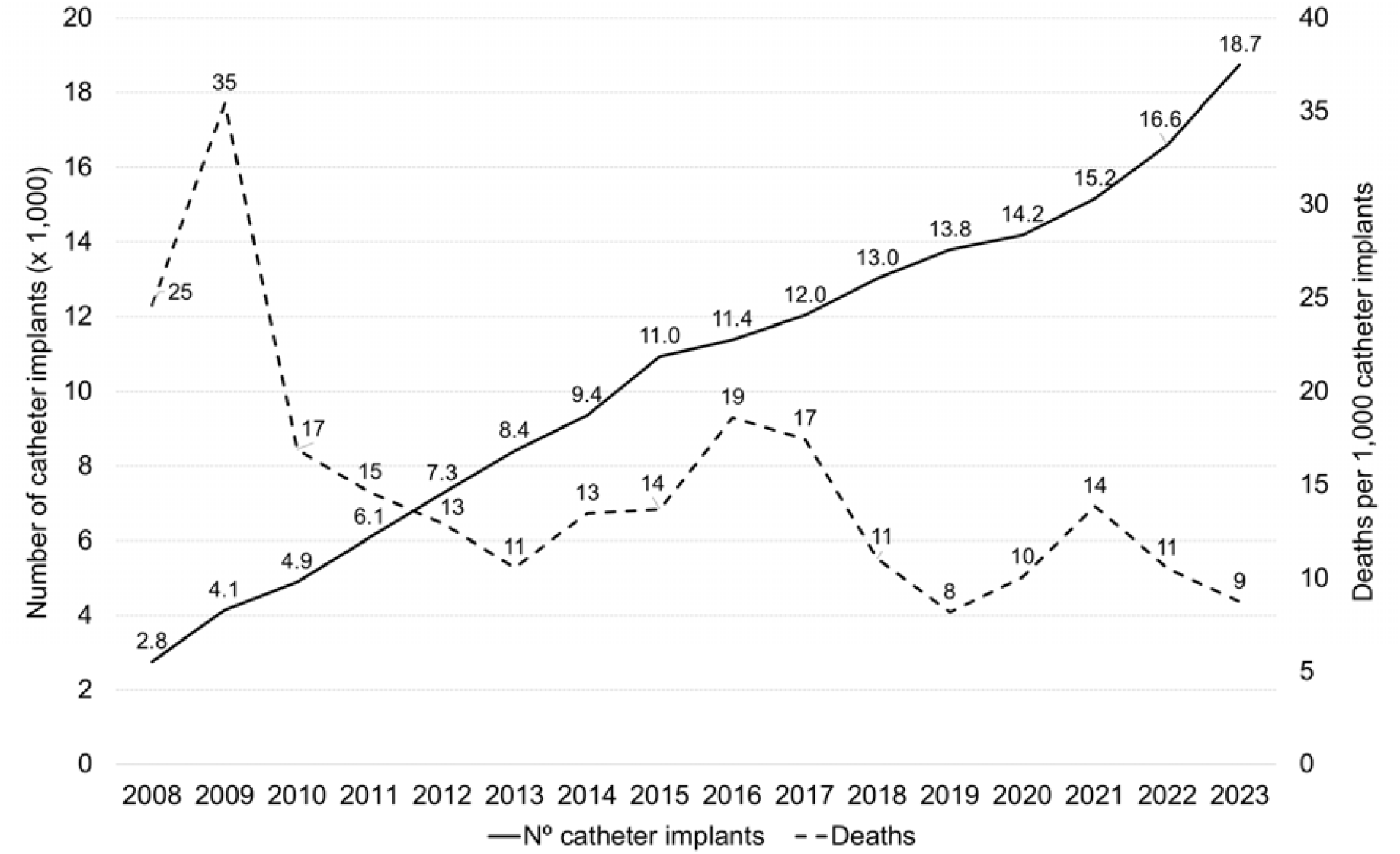
Lethality over the years

Table 5 shows the number of deaths attributable to various diagnoses in patients following long-term catheter implantation, with nearly 300 attributed to lymphatic, hematopoietic, and related tissue neoplasms, the leading cause of death. Fatalities were concentrated in a few oncologic categories, with a steep drop-off in rarer diagnoses. Among 122,271 oncologic procedures, 1,187 deaths occurred (10 per 1,000 implants). Mortality rates varied from 6 to 25 per 1,000—highest for respiratory system neoplasms, lowest for digestive tract and breast cancers—highlighting distinct risk profiles among tumor types.

**Table 5.** Distribution of deaths and lethality rates by primary diagnosis among patients undergoing long-term catheter implantation in Brazil (2008-2023).

| DIAGNOSIS | NUMBER OF DEATHS | NUMBER OF PROCEDURES | DEATHS PER 1000 IMPLANTS | P-VALUE |
| --- | --- | --- | --- | --- |
| Neoplasms of lymphatic, haematopoietic and related tissues | 300 | 24 676 | 12 | REFERENCE |
| Digestive tract neoplasms | 227 | 37 678 | 6 | <0.001 |
| Breast neoplasms | 186 | 25 901 | 7 | <0.001 |
| Neoplasms of the respiratory system and intrathoracic organs | 121 | 4 856 | 25 | <0.001 |
| Female reproductive system neoplasms | 117 | 6 156 | 19 | <0.001 |
| Neoplasms of the liver, pancreas and bile ducts | 69 | 6 233 | 11 | 0.483 |
| Head and neck neoplasms | 57 | 4 247 | 13 | 0.494 |
| Bone, cartilage, connective tissue and soft tissue neoplasms | 34 | 5 330 | 6 | <0.001 |
| Urinary system neoplasms | 29 | 1 649 | 18 | 0.058 |
| Nervous system neoplasms | 16 | 2 417 | 7 | 0.018 |
| Skin neoplasms, mesothelioma and Kaposi's sarcoma | 11 | 1 158 | 9 | 0.422 |
| Male reproductive system neoplasm | 10 | 1 019 | 10 | 0.505 |
| Adrenal and other endocrine gland neoplasms | 10 | 951 | 11 | 0.652 |

Regarding the financial aspect, the total amount reimbursed by the PUBLIC system for these procedures was USD 19,478,056.93, while the average reimbursement per hospitalization for the implantation of long-term catheters was USD 115.33.

## DISCUSSION

This retrospective cross-sectional study analyzed 168,869 long-term catheter implantations performed across Brazil between 2008 and 2023, covering about 160 million people. Data were obtained from DATASUS—the national public health database that records all reimbursed procedures—and population statistics from the Brazilian Institute of Geography and Statistics (IBGE). DATASUS provides anonymized information on patients treated within the public system, including demographics, hospital admissions, length of stay, costs, and procedure details. This integration allowed precise demographic stratification and standardized rate calculations, resulting in the first and most comprehensive nationwide evaluation of long-term catheter implantation practices in Brazil.

We observed a steady and progressive increase in the number of procedures over the years, likely driven by the rising incidence of oncological diseases, the wider availability of vascular access devices, and significant improvements in healthcare infrastructure that facilitate their implantation [18].

The data highlight a consistent growth in both absolute numbers and the relative contribution of successive years to the total volume of implants, indicating sustained national expansion and consolidation in the use of long-term catheters as part of modern oncologic care across Brazil.

Publications on the use of venous access for chemotherapy in developed countries indicate that, in the United States alone, more than 500,000 long-term catheters are implanted annually [19]. Surveys conducted in European countries also report high utilization rates of these devices. According to NHS data, in the United Kingdom, approximately 150,000 oncology patients use long-term catheters for chemotherapy each year [20]. However, there is a lack of data on catheter implantation rates in low- and middle-income countries, highlighting a gap in knowledge regarding this type of medical care and the disparities present in these settings.

The demographic profile of the sample showed a predominance of female patients, probably secondary to the higher prevalence of malignant neoplasms more common in women, such as breast cancer (the most common malignant neoplasm in the country) and colorectal cancer (the second most common malignant neoplasm among women and the third in Brazil) [21]. Furthermore, the most common neoplasm in men, prostate cancer [21], rarely requires long-term systemic chemotherapy, as it is generally managed through surgery, radiotherapy, or hormonal therapy, with chemotherapy reserved mainly for advanced and palliative cases [22]. Consequently, the use of long-term access in men with cancer is more often associated with less prevalent malignancies, which contributes to a lower number of catheters in men.

The distribution of diagnoses associated with long-term venous catheters in Brazil reveals that oncologic diseases are the main indications for implantation. Digestive tract neoplasms accounted for the largest share (22.3%), followed by breast (15.3%) and hematologic malignancies (14.6%), all of which commonly require prolonged chemotherapy. Venous disorders (19.4%) probably reflect non-oncologic uses such as long-term antibiotic therapy or parenteral nutrition. Breast câncer treatment frequently involves chemotherapy at multiple stages, making long-term catheters, particularly fully implantable ones, essential for safe and durable venous access [23,24]. In colorectal cancer, chemotherapy is typically administered in advanced stages, and the high frequency of late diagnoses in Brazil, due to limited national screening programs, justifies the increased need for these devices [25] [26,27].

According to the National Cancer Institute (INCA), 704,000 new cases of cancer are expected between 2023-2025, with approximately 65% occurring in the South and Southeast, further emphasizing the demand for long-term venous access in these two regions [21].

Most patients who underwent long-term catheter implantation had brief hospital stays, with 74.3% discharged within one day, reflecting the procedure’s efficiency and low complication rate. The total public expenditure for these implantations reached approximately US$19.5 million, resulting in an average reimbursement of US$115.33 per hospitalization. In comparison, a Swedish study assessing the costs related to catheter implantation reported a significantly higher average cost of US$371.76, while also reinforcing the superior cost-effectiveness of this method compared to other approaches for chemotherapy administration [28].

This discrepancy may reflect differences in healthcare systems, material costs, and medical fees. However, it also underscores the substantially lower cost of long-term catheter implantation in Brazil, which may contribute to reducing overall cancer treatment expenses and optimizing healthcare resource allocation.

The distribution of deaths following long-term catheter implantation shows that most fatalities occurred in oncologic conditions, especially hematopoietic, lymphatic, and digestive tract neoplasms. This pattern reflects the main use of long-term catheters in cancer care— chemotherapy, parenteral nutrition, and repeated infusions—in patients already at high risk. The predominance of hematologic malignancies corresponds to aggressive forms like leukaemia and lymphoma, which require intensive therapy and carry high mortality. Digestive tract cancers also appear prominently, reflecting their late diagnosis and poor prognosis in Brazil. Venous disorders among the leading causes may indicate that vascular or thrombotic complications contribute to mortality [29]. These data represent a high-risk cohort, not the general cancer population, as patients requiring implanted catheters often have advanced disease [30].

Regional disparities may also play a role, since specialized centers are concentrated in the Southeast and South. Overall, these findings highlight the strong association between long-term catheter use and severe, high-mortality oncologic conditions [31].

## LIMITATIONS

This analysis has limitations that must be considered when interpreting the results. The study only included procedures carried out in the public health system, which, despite accounting for 70% of health care [17], does not reflect the reality of all medical care in the country, since data from the private sector were not evaluated. In addition, we did not have access to patients’ medical records, since this information was extracted from an official federal government database, which is filled in using records made by health institutions, which can be subject to registration errors. An example of this is the high prevalence of ICD I87 (Other vein disorders) in long-term catheter implants, which may not accurately reflect the clinical indication for the use of the device. Finally, among the patients who died, it was not possible to define the impact exclusively associated with the procedure (catheter implantation) on the observed mortality.

## CONCLUSIONS

Based on a total of 168,869 chemotherapy catheters implanted from 2008 to 2023 in the Brazilian public health system, we conclude that most recipients are adult women, (aged 30– 64 years) with cancer (colorectal, breast, and hematolymphoid neoplasms). The associated mortality rate is low but not null (1.3%) and the cost estimated per procedure is US$115.33.

## Supporting information

TITLE PAGE

## Data Availability

All data produced in the present study are available upon reasonable request to the authors

https://datasus.saude.gov.br/

