## Supplementary material for "Epidemiology of Long-Term Catheter Implantation for Chemotherapy in the Public Health System in Brazil: A 16-Year Analysis of 168,869 Cases": TITLE PAGE

**Affiliations:**

Affiliation: Hospital Israelita Albert Einstein – Vascular and Endovascular Surgery

Division: São Paulo, SP, Brazil

Postal Address: Av Albert Einstein, 627, Bloco A1, Sala 423, ZIP Code 05652-900. Morumbi, São Paulo – SP, Brazil

Phone Number: +55 11 2151 - 5423

**ORCID:**

Arthur Soares Freitas e Silva: 0009-0001-7772-4125

Nahmam Jarouche: 0009-0000-5752-4268

Carolina Carvalho Jansen Sorbello: 0009-0004-1229-5130

Bruno Jeronimo Ponte: 0000-0002-7272-7423

Marcelo Fiorelli Alexandrino da Silva: 0000-0002-0714-5291

Marcelo Passos Teivelis: 0000-0002-3648-6773

Nelson Wolosker: 0000-0003-1991-3507

**Conflicts of Interest and Source of Funding:** The authors declare no conflicts of interest. This research did not receive any specific grants from funding agencies in the public, commercial, or not-for-profit sectors.
